# OphthUS-GPT: Multimodal AI for Automated Reporting in Ophthalmic B-Scan Ultrasound

**DOI:** 10.1101/2025.03.03.25323237

**Authors:** Fan Gan, Lei Chen, Weiguo Qin, Qingluan Han, Xin Long, Huimin Fan, Xinyuan Li, Hanzhu Yu, Junhan Zhang, Nuo Xu, Jianxun Cheng, Jian Cao, Kangcheng Liu, Yinan Shao, Xiaonan Li, Qi Wan, Teng Liu, Zhipeng You

## Abstract

**IMPORTANCE:** The rapid advancement of AI in ophthalmology is transforming diagnostics, especially in resource-limited settings. The shortage of ophthalmologists and lack of standardized reporting creates an urgent need for AI systems capable of automated reporting and interactive decision support.

**OBJECTIVE:** To develop OphthUS-GPT, a multimodal AI system integrating BLIP and DeepSeek models for automated report generation and clinical decision support from ophthalmic B-scan ultrasound images.

**DESIGN, SETTING, AND PARTICIPANTS:** This retrospective study at the Affiliated Eye Hospital of Jiangxi Medical College collected B-scan ultrasound reports between 2017-2024, including 54,696 images and 9,392 reports from 31,943 patients (mean age 49.14±0.124 years, 50.15% male).

**MAIN OUTCOMES AND MEASURES:** Evaluation included two components: diagnostic report generation and question-answering system assessment. Report generation was evaluated using text metrics (ROUGE-L, CIDEr), disease classification metrics (accuracy, sensitivity, specificity, precision, F1 score), and ophthalmologist ratings for accuracy and completeness. The question-answering system was assessed by ophthalmologists rating answers on accuracy, completeness, potential harm, and satisfaction.

**RESULTS:** OphthUS-GPT achieved ROUGE-L and CIDEr scores of 0.6131 and 0.9818 in report generation. For common conditions, accuracy exceeded 90% with precision >70%. Expert assessment showed >90% of reports scored ≥ 3/5 for correctness and 96% for completeness. The DeepSeek-R1-Distill-Llama-8B (DeepSeek) question-answering component performed comparably to GPT4o and OpenAI-o1, outperforming other models.

**CONCLUSIONS AND RELEVANC:** OphthUS-GPT demonstrated excellent performance in automatic report generation and intelligent Q&A, offering a novel solution for ophthalmic ultrasound interpretation and clinical decision support.

## Introduction

Deep learning-driven artificial intelligence (AI) technologies are driving the Fourth Industrial Revolution.^1,2^ In ophthalmology, the combination of intelligent fundus photography devices and telemedicine has greatly improved the quality of community-level healthcare services, making ophthalmic disease screening more efficient and convenient.^3–5^ However, the lack of ophthalmologists and specialized equipment in primary care limits telemedicine implementation and the allocation of high-quality medical resources.^6,7^ Additionally, with improving living standards and an aging population, the prevalence of cataracts and diabetic retinopathy is increasing, especially among the elderly, where cataracts often coexist with other ocular conditions.^8–12^

As a non-invasive and cost-effective diagnostic tool, ultrasonography has been widely adopted in primary care institutions.^13–15^ It enables the evaluation of the posterior segment in cases of media opacity and plays a vital role in postoperative retinal monitoring.^16,17^ While primary care personnel can perform ultrasound examinations independently, even without guidance from specialized radiologists.^18–21^ They often lack the expertise to write diagnostic reports, and their subjective interpretations can vary due to differences in experience, potentially affecting telemedicine workflows.^22,23^ Additionally, in tertiary hospitals, drafting diagnostic reports remains time-consuming and significantly adds to the workload of physicians.^24,25^

Since 2022, advancements like the Bootstrapping Language-Image Pre-training (BLIP)^26^ model has successfully integrated visual information with natural language processing, enabling applications like automated MRI images description and Fundus fluorescein angiography (FFA) reports generation.^26–28^ Meanwhile, the emergence of large language models (LLMs) like DeepSeek-R1-Distill-Llama-8B (Deepseek)^29^ has driven a paradigm shift toward human-machine interactive learning in data-driven approaches.^30,31^ In the medical domain, LLMs have effectively assisted physicians in interpreting pathological tissue slides and chest X-rays through question-and-answer interactions.^32,33^

To build on these advancements, this study proposes a two-stage artificial intelligence system named OphthUS-GPT. The system utilizes the BLIP model to analyze ultrasound images and generate diagnostic reports that meet medical standards. Additionally, it incorporates the Deepseek model to provide multi-turn intelligent dialogue support for explaining reports to both patients and physicians. By deploying the system in the cloud, OphthUS-GPT aims to enable real-time telemedicine collaboration between higher-tier hospitals and primary healthcare institutions. The system is designed to standardize diagnostic report content, alleviate the shortage of clinical resources, and improve the efficiency of telemedicine collaboration. The workflow and demonstration of this study is illustrated in **Figure 1**.

**Figrue 1.**
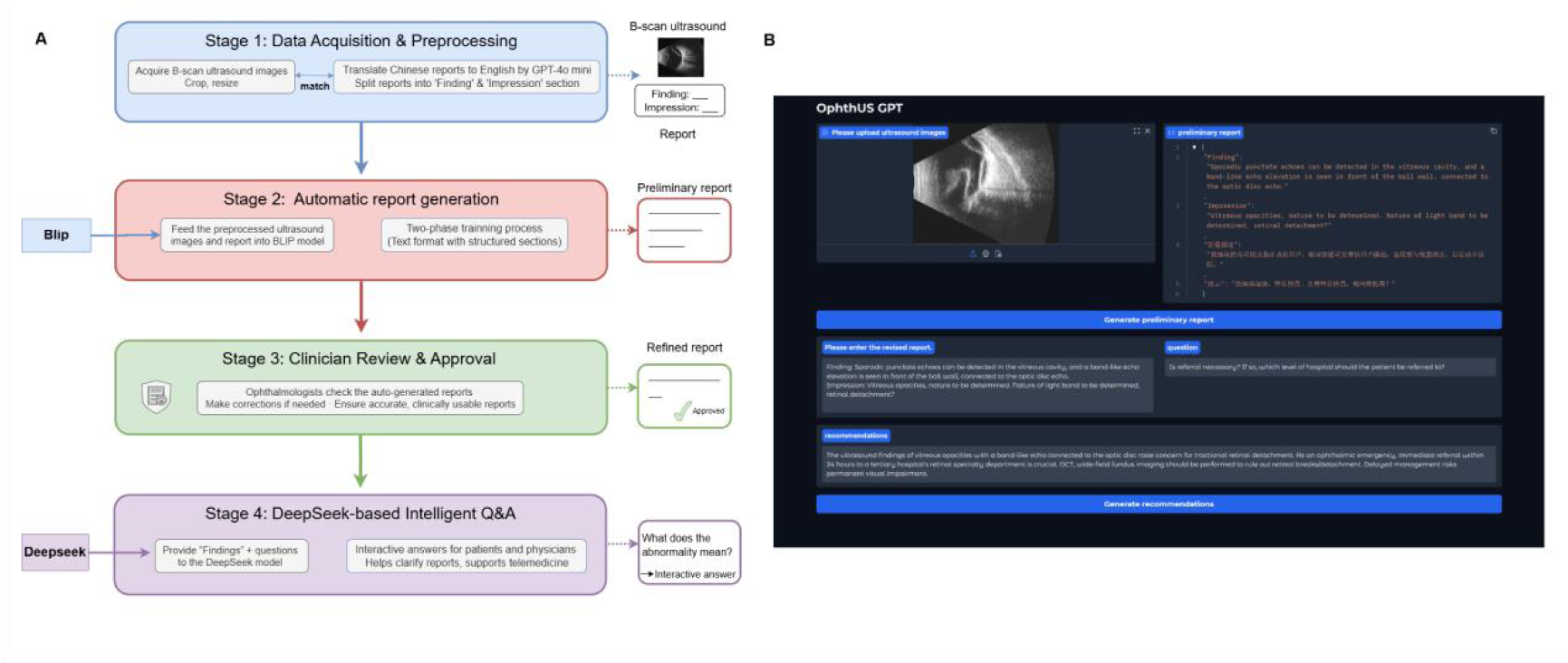
Overview of the workflow and demonstration of this study. A, Flow chart of the entire experiment. B, Demonstration of the integrated interface within OphthUS-GPT.

## Method

### Dataset

This study adhered to the principles of the Declaration of Helsinki and was approved by the Ethics Committee of the Affiliated Eye Hospital of Jiangxi Medical College, Nanchang University (approval number: YLP20220802). Given the retrospective nature of the analysis, the requirement for informed consent was waived. We retrospectively collected diagnostic reports of ophthalmic B-scan ultrasound examinations at the Affiliated Eye Hospital of Nanchang University between June 2017 and March 2024. Each report included ophthalmic B-scan ultrasound images of the right and left eyes, along with a free-text description of the findings and the ophthalmologist’s impression. All image data were acquired using the MD-2400S ultrasonography device. To protect patient privacy, all identifiable information was anonymized.

### Data Preprocessing

The original size of all images was at a resolution of 720×576 pixels, we utilized a standardized cropping procedure to remove extraneous areas, specifically cropping 70 pixels from the left and top edges, and 70 pixels from the right edge and 55 pixels from the bottom edge, to better isolate the region of interest. The cropped images were subsequently resized to a uniform dimension of 320 × 320 pixels, ensuring consistent input for the deep learning model.

The diagnostic reports were originally written in Chinese. To maintain a single-language framework, we used GPT-4o-mini for translation into English, facilitating uniform text tokenization due to the English-based pretraining of our language model component. On the textual side, each training sample’s diagnostic report was parsed into two fields: (i) “Findings”, which provide an objective record of the imaging features observed during the ultrasound examination, including the morphology, size, location, and internal echogenic characteristics of any lesions, and (ii) “Impression”, which represents a preliminary diagnostic conclusion or potential diagnostic suggestion, derived from the findings and integrated with clinical information.

### Development of OphthUS-GPT

Our report generation framework employed the BLIP model^26^ as the backbone. BLIP consists of a Visual Transformer encoder and a bidirectional text encoder-decoder based on BERT^34^.This enables efficient cross-modal representation learning, making it suitable for our ocular ultrasound image-to-report generation task.

During the report generation phase, we fine-tuned the BLIP model using a two-phase training strategy with pretrained weights. In Phase 1, the training objective was limited to generating the “Findings” section. Each image-report pair was formatted to help the model learn to produce image descriptions that began with the prefix “Findings:”. This phase involved training for 5 epochs, allowing the model to effectively capture key ocular lesions without overfitting to the training data. In Phase 2, the training objective was extended to include both the “Findings” and “Impression” sections. This phase spanned 20 epochs, during which we observed consistent improvements in model performance compared to a single-phase training approach. Model optimization was carried out using standard techniques, and the best checkpoint was selected based on the BERTScore^35^ metric. Training was conducted on an A800 GPU, with specific hyperparameter details provided in **eTable 1 in Supplement 1**.

During the question and answering phase, we developed 10 clinically relevant questions, 5 from the patient’s perspective and 5 from the ophthalmologist’s, drawing on clinical expertise and prior research.^28^ The generated “findings” and corresponding questions were input into the DeepSeek. Guided by a tailored prompt, DeepSeek provided answers without requiring domain-specific fine-tuning, thereby improving the reports’ accessibility and effectiveness. The questions and prompt are detailed in **eTable 2 in Supplement 1**.

### Performance Evaluation

To evaluate the performance of the generated reports, we utilized a combined approach that included both quantitative evaluation and human assessment. The quantitative evaluation involved language-based metrics and disease classification metrics. We employed BLEU^36^, ROUGE^37^, and BERTScore to assess the language similarity and coherence of the generated reports. Furthermore, we extracted key disease categories (e.g., vitreous hemorrhage) from the reports through keyword matching and subsequently calculated classification performance metrics, such as accuracy, sensitivity, specificity, precision, and F1 score. This approach captures sparse yet critical disease-related keywords, thereby overcoming the limitations of traditional metrics. Additionally, we randomly selected 48 test samples for independent evaluation by three ophthalmologists. These experts rated the “accuracy” and “completeness” of the reports on a five-point scale, with the specific scoring criteria detailed in **eTable 3 in Supplement 1**.

To evaluate the performance of OphthUS-GPT in disease classification tasks more comprehensively, we conducted a comparative experiment against mainstream large language models. A total of 420 test samples were selected, and multiple mainstream large language models, namely GPT-4 Turbo^38^, GPT-4o^39^, Claude 3.5 sonnet (Claude)^40^, and WiNGPT2-Llama-3-8B-Chat(WiNGPT2)^41^ were evaluated under 1-shot, 3-shot, and 5-shot learning settings. Notably, due to significantly lower performance in the 1-shot and 3-shot settings, the 5-shot experiments for WiNGPT2 were omitted to optimize computational resources.

To evaluate the performance of the question-answering system, we randomly selected 48 samples from the test set (3 samples per category). For each case, six questions were selected from a pre-designed comprehensive question set, including three patient-related questions and three physician-related questions. These questions, along with the generated reports, were input into the following large language models, including DeepSeek, GPT-4o, GPT-4 Turbo, Claude, OpenAI-o1^42^, and WiNGPT2. Subsequently, three ophthalmologists evaluated the generated responses based on a 5-point scoring standard for question-answering, assessing the results across four dimensions: “accuracy”, “completeness”, “potential harm” and “satisfaction”. The scoring criteria are detailed in **eTable 4 in Supplement 1**. Through this process, we assessed and compared the performance of DeepSeek and other large language models in answering common questions about ophthalmic diseases.

### Statistical Analysis

All statistical analyses were performed in Python 3.8.5. with p-values below 0.05 considered statistically significant. Fleiss’ Kappa was employed to evaluate inter-rater agreement among three ophthalmologists, with a Kappa value ≥0.6 indicating strong consistency. When Kappa values were ≥ 0.6, the median score from the three ophthalmologists was taken as the final evaluation score for each question-answer pair. The Kruskal-Wallis H test was used to compare evaluation scores across six models, followed by Dunn’s test with Holm correction for pairwise comparisons. If the Kappa value was <0.6, reflecting insufficient agreement, a panel of retinal disease experts determined the final score.

## Results

### Data Characteristics

The final dataset consisted of 54,696 ophthalmic B-scan ultrasound images and 9,392 reports from 31,943 patients. To ensure external validation, we divided the dataset into training, validation, and testing subsets using temporal splits. Among these, 42,688 images (78.05%, 21,344 reports) captured from June 2017 to December 2022 were used for training, 6,004 images (10.98%, 3,002 reports) from 2023 were used for validation, and 6,004 images (10.98%, 3,002 reports) from 2024 were used for testing. The eye conditions extracted from the original ophthalmic B-scan ultrasound reports primarily encompass a range of common ocular pathologies, including vitreous opacities, posterior vitreous detachment, posterior scleral staphyloma, retinal detachment, and vitreous hemorrhage. The average age of the patients was 49.14 ± 0.124 years, with 16,021 males (50.15%). The detailed characteristics of the dataset are shown in **Table 1**.

**Table 1.** Clinical Characteristics of the Dataset.

| Characteristic | Total | Train | Validation | Test |
| --- | --- | --- | --- | --- |
| Population |  |  |  |  |
| No. | 31943 | 22509 | 4714 | 4720 |
| Age, mean±SD (y) | 49.14±0.124 | 48.74±0.149 | 49.83±0.319 | 49.69±0.321 |
| Gender, n (%) |  |  |  |  |
| Male | 16021 (50.15) | 10992(51.2) | 2500(53) | 2529(53.6) |
| Female | 15922 (49.85) | 11517(48.8) | 2214(47) | 2191(46.4) |
| Ultrasonography Images |  |  |  |  |
| No. | 54696 | 42688 | 6004 | 6004 |
| Conditions |  |  |  |  |
| Vitreous Opacities | 35730 | 29065 | 3373 | 3292 |
| PVD | 18594 | 14792 | 1947 | 1855 |
| NSA | 8705 | 6988 | 836 | 881 |
| PSS | 8701 | 5846 | 1448 | 1407 |
| PSOTS | 7372 | 4688 | 1337 | 1347 |
| Intraocular Lens | 5625 | 4038 | 772 | 815 |
| LBNPI | 3881 | 2323 | 794 | 764 |
| Retinal Detachment | 3266 | 1963 | 671 | 632 |
| Cataract | 2050 | 1885 | 95 | 70 |
| PSORS | 2425 | 1628 | 384 | 413 |
| Complete PVD | 3118 | 1613 | 758 | 747 |
| Vitreous Hemorrhage | 2067 | 1135 | 466 | 466 |
| Asteroid Hyalosis | 874 | 551 | 157 | 166 |
| Choroidal Detachment | 285 | 192 | 46 | 47 |
| Phthisis Bulbi | 296 | 166 | 66 | 64 |
| Lens Dislocation | 248 | 163 | 41 | 44 |

Abbreviations: PVD: Posterior Vitreous Detachment; NSA: No Significant Abnormalities; PSS: Posterior Scleral Staphyloma; PSOTS: Post-Silicone Oil Tamponade Surgery; LBNPI: Light Band Nature Pending Investigation; PSORS: Post-Silicone Oil Removal Surgery

### Reports Generation Performance

We objectively evaluated the report generation performance of our system in terms of text generation quality and disease classification accuracy. Through a series of ablation studies and comparisons with the R2Gen series models, we systematically analyzed the impact of various training strategies and model architectures on report generation performance. In terms of text generation quality, all models demonstrated excellent performance on the test set. Among them, the Finding5epoch+Impression model achieved the best results, with ROUGE-L and CIDEr scores of 0.6131 and 0.9818, respectively, indicating high semantic consistency and superior text generation quality. The performance metrics for text generation are presented in **Table 2**.

**Table 2.** Text generation performance of OphthUS-GPT in the test set.

| Model | BLEU-1 | BLEU-2 | BLEU-3 | BLEU-4 | ROUGE-L | BERT Score | CIDEr |
| --- | --- | --- | --- | --- | --- | --- | --- |
| Finding | 0.4695 | 0.3958 | 0.3403 | 0.2874 | 0.4840 | 0.9300 | 0.8542 |
| Finding+Impression | 0.5387 | 0.4323 | 0.3660 | 0.3072 | 0.5612 | 0.9262 | 0.9109 |
| Finding5epoch+Impression | 0.5739 | 0.4793 | 0.4151 | 0.3552 | 0.6131 | 0.9329 | 0.9818 |
| R2Gen | 0.4894 | 0.3983 | 0.3393 | 0.2850 | 0.5534 | 0.9266 | 0.9446 |
| R2Gen+CMN | 0.5552 | 0.4637 | 0.4006 | 0.3418 | 0.6050 | 0.9313 | 0.9461 |

Regarding ophthalmic disease diagnosis, our model demonstrated exceptional performance in detecting a wide range of ocular conditions, with diagnostic accuracy exceeding 80% for most diseases. Notably, for common conditions such as vitreous opacities, retinal detachment, posterior scleral staphyloma, and cataracts, the accuracy exceeded 90%, with precision greater than 70%. For rarer conditions, including choroidal detachment and phthisis bulbi, the diagnostic accuracy reached 0.9893 and 0.9962, with sensitivity values of 0.7377 and 0.8070, respectively, and specificity exceeding 99%. Additionally, the model exhibited an accuracy of over 96% for diagnosing postoperative silicone oil removal status, with a sensitivity of 99%. For lens dislocation diagnosis, the model achieved accuracy, specificity, and sensitivity values of 0.9930, 0.9998, and 0.7353, respectively. However, the model’s precision for vitreous hemorrhage and asteroid hyalosis was relatively lower, at 0.6716 and 0.5684, respectively. This is likely due to the imbalanced distribution of data, with fewer samples available for these conditions, which may have hindered the model’s ability to effectively learn and differentiate their features. The performance metrics for disease classification are presented in **Table 3**.

**Table 3.** Classification performance of OphthUS-GPT in the test set.

| Diagnosis | Accuracy | Specificity | Precision | Sensitivity | F1 Score |
| --- | --- | --- | --- | --- | --- |
| Vitreous Opacities | 0.9360 | 0.8997 | 0.9193 | 0.9668 | 0.9424 |
| PVD | 0.8226 | 0.9060 | 0.8083 | 0.7589 | 0.7828 |
| NSA | 0.9357 | 0.9486 | 0.6440 | 0.8213 | 0.7219 |
| PSS | 0.9314 | 0.9662 | 0.8719 | 0.8089 | 0.8392 |
| PSOTS | 0.9652 | 0.9578 | 0.8515 | 0.9957 | 0.9180 |
| Intraocular Lens | 0.8023 | 0.7952 | 0.7321 | 0.8628 | 0.7921 |
| LBNPI | 0.9156 | 0.9355 | 0.8275 | 0.7172 | 0.6079 |
| Retinal Detachment | 0.9385 | 0.9664 | 0.7147 | 0.7047 | 0.7097 |
| Cataract | 0.9880 | 0.9965 | 0.7474 | 0.7250 | 0.7360 |
| PSORS | 0.9742 | 0.9900 | 0.8440 | 0.7537 | 0.7963 |
| Complete PVD | 0.9757 | 0.9898 | 0.8866 | 0.8295 | 0.8571 |
| Vitreous Hemorrhage | 0.9459 | 0.9758 | 0.6716 | 0.5892 | 0.6277 |
| Asteroid Hyalosis | 0.9777 | 0.9827 | 0.5684 | 0.8012 | 0.6650 |
| Choroidal Detachment | 0.9893 | 0.9978 | 0.8971 | 0.7377 | 0.8097 |
| Phthisis Bulbi | 0.9962 | 0.9980 | 0.7931 | 0.8070 | 0.8000 |
| Lens Dislocation | 0.9930 | 0.9998 | 0.7855 | 0.7353 | 0.7596 |

Abbreviations: PVD: Posterior Vitreous Detachment; NSA: No Significant Abnormalities; PSS: Posterior Scleral Staphyloma; PSOTS: Post-Silicone Oil Tamponade Surgery; LBNPI: Light Band Nature Pending Investigation; PSORS: Post-Silicone Oil Removal Surgery

We also conducted a comparative evaluation against contemporary LLMs. While our multimodal model was comprehensively evaluated on the full test set, these models were tested on only 420 samples under few-shot learning conditions. Despite their capabilities, these models showed significant limitations in this restricted learning scenario. Our approach consistently achieved more balanced and comprehensive diagnostic capabilities, particularly for complex conditions. For instance, the F1 score for Post-Silicone Oil Tamponade Surgery (PSOTS) reaches 0.9180, posterior vitreous detachment achieves 0.8571. Furthermore, our model exhibits remarkable advantages in specificity (e.g., choroidal detachment: 99.78%) and precision (e.g., vitreous opacity: 91.93%). These results strongly validate the significant potential and irreplaceable advantages of multimodal data fusion in enhancing diagnostic accuracy and robustness. The details are shown in **Table 3** and **eTable 5 in Supplement 1**.

Three ophthalmologists manually evaluated the generated reports in terms of “accuracy” and “completeness.” The Fleiss’ kappa values demonstrated high inter-rater agreement, with a kappa value of 0.923 for accuracy and 0.937 for completeness (**eTable 6 in Supplement 1**). Over 90% of the reports achieved a score of 3 or higher for correctness, and 96% of the reports achieved a score of 3 or higher for completeness. Reports scoring 3 or higher were deemed potentially usable for clinical purposes after appropriate supplementation and modification. Less than 2% of the reports were deemed unsuitable for clinical use due to complete inconsistency with actual conditions or omission of critical information. This distribution is shown in **Figure 2**. These errors were primarily attributed to inaccurate descriptions of ultrasound features, misidentification of complex lesions, and false negatives resulting from unclear images. Examples of generated reports at different levels are provided in **eFigure 1 in Supplement 1**.

**Figrue 2.**
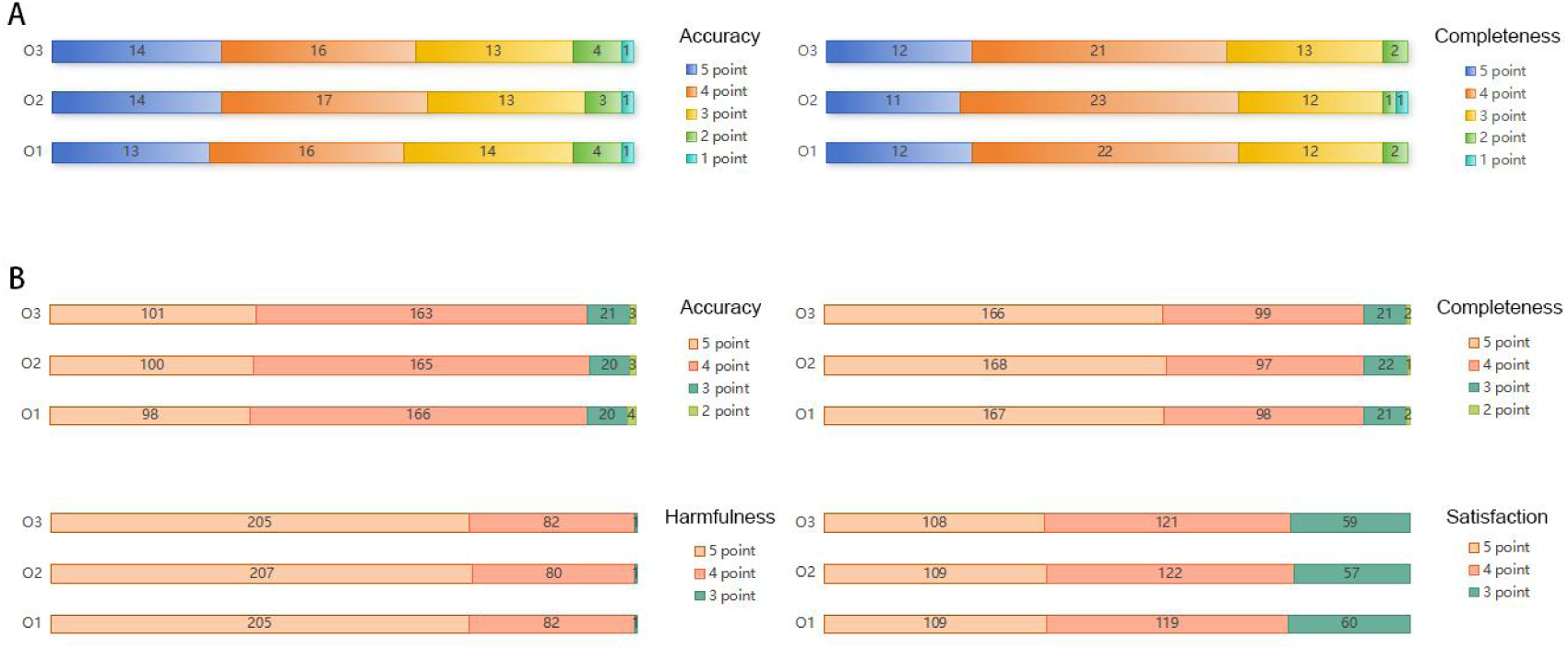
Performance evaluation of OphthUS-GPT by ophthalmologists. A, Evaluation of report generation capabilities. B, Evaluation of question answering capabilities. O, Ophthalmologist.

### Question-Answering System Performance

We assessed the inter-rater reliability of three ophthalmologists in evaluating the quality of model responses using the Kappa consistency test. Results showed that the Kappa values for accuracy, completeness, potential harm, and satisfaction dimensions all exceeded 0.95, indicating high reliability of the scoring process (see **eTable 7 in Supplement 1**). The Kruskal-Wallis test revealed no statistically significant differences among six large language models in the completeness (H=2.938, p=0.710) and potential harm (H=6.510, p=0.260) dimensions. Notably, the potential harm scores were generally above 3, and completeness scores exceeded 2 across all models, suggesting that the responses contained no significant omissions and posed no serious health risks. However, significant differences were observed among models in the accuracy (H=15.352, p=0.009) and satisfaction (H=34.640, p<0.001) dimensions. Post hoc analysis using Dunn’s test with Holm correction demonstrated that, in terms of accuracy, the DeepSeek model significantly outperformed the Claude model (p=0.011), with the majority of its samples scoring above 3 and only a few scoring 2. For satisfaction, there were no statistically significant differences between DeepSeek, GPT4o, and OpenAI-o1, with all three models scoring above 3. However, DeepSeek showed a significant advantage over the remaining models (p<0.005), as detailed in **Figure 2**, **eFigure 2 in Supplement 1** and **eTable 8 in Supplement 1**. These findings indicate that DeepSeek performs exceptionally well across accuracy, completeness, potential harm, and satisfaction dimensions, and its satisfaction scores are comparable to those of the most advanced models currently available. The evaluation examples of the relevant Q&A pairs are provided in **eTable 9 in Supplement 1**.

## Discussion

This study developed a two-stage AI system named OphthUS-GPT. In the first stage, the system automatically analyzed ultrasound images using the BLIP model and generated diagnostic reports that met medical standards. These reports were then reviewed and confirmed by ophthalmologists. In the second stage, the system employed the DeepSeek model to conduct interactive question-and-answer sessions based on the report content. Through comprehensive automated and manual evaluations, the system has demonstrated reliable and satisfactory performance.

The rapid advance of AI in the field of ophthalmology is swiftly transforming traditional diagnostic approaches.^43,44^ While previous single-modality models achieved >80% diagnostic accuracy with ophthalmic B-scan ultrasound images, they lacked detailed feature descriptions.^45–47^ Our BLIP-based approach with progressive training achieves >90% accuracy while generating comprehensive reports rather than mere diagnostic labels. Furthermore, by inputting the generated diagnostic description texts into mainstream large language models, we demonstrated the superiority of multimodal models over single-modality text-based models in diagnostic performance.^48^ Crucially, our system not only outputs diagnostic categories but also automates the generation of high-quality diagnostic reports. This automation effectively alleviates the workload associated with image interpretation and report writing for clinicians, thereby significantly improving diagnostic efficiency. Additionally, the standardized report format helps to reduce diagnostic inconsistencies caused by individual experience differences, providing high-quality diagnostic support for primary and telemedicine settings.^49–51^

In previous studies, Wang et al^52^ developed an ophthalmic ultrasound diagnostic report generation system based on cross-modal deep learning. However, our study not only achieved automated diagnostic report generation but also utilized DeepSeek to construct a professional question-answering system for the interpretation of ophthalmic ultrasound diagnostic reports. By leveraging DeepSeek, a specialized question-answering system was constructed, successfully integrating the diagnostic report module with the clinical decision-making support module. This integration enabled a fully automated workflow from diagnostic report generation to clinical decision-making. Zhao et al^53^ and Chen et al^28,54^ have respectively developed automated report generation and question-answering systems based on slit-lamp images, FFA, and Indocyanine green angiography (ICGA). These systems utilized models such as Llama 2 and GPT-4 for question-answering system construction, but these models have significant drawbacks, including insufficient adaptation to medical knowledge, poor interpretability of diagnostic logic, and high deployment costs. In this study, we introduce a novel approach leveraging the lightweight architecture of the DeepSeek model, which not only ensures high performance but also significantly reduces the actual deployment costs in clinical settings. To further evaluate the performance of DeepSeek, we compared it with five mainstream large language models, including Claude, GPT-4o, GPT-4-turbo, OpenAI-o1, and WiNGPT2. The results demonstrate that DeepSeek exhibits superior performance in multiple core dimensions, including accuracy, completeness, potential harm, and satisfaction, with performance comparable to OpenAI-o1 and GPT-4o. This finding not only highlights the advantages of DeepSeek in interpreting ophthalmic ultrasound reports but also provides valuable insights for the selection and optimization of AI-assisted systems in ophthalmology.

However, our study not only achieved automated diagnostic report generation but also utilized DeepSeek to construct a professional question-answering system for the interpretation of ophthalmic ultrasound diagnostic reports. By leveraging DeepSeek, we successfully constructed a specialized question-answering system, integrating the diagnostic report module with the clinical decision-making support module. This integration enabled a fully automated workflow from diagnostic report generation to clinical decision-making.

### Limitations

Certainly, our study also has certain limitations. First, while multimodal medical imaging is a growing trend in AI development, our model is limited to ophthalmic B-scan images. Future work should explore multimodal imaging models. Second, the model has only been tested on time-split data and lacks validation on independent primary care datasets. Additionally, low-quality images may increase misdiagnosis risks, highlighting the need for future efforts in image enhancement and model optimization.

## Conclusions

In summary, this study combines the BLIP and DeepSeek models to develop OphthUS-GPT, a system that automates reporting and enables intelligent Q&A based on ophthalmic B-scan images. It offers a novel solution for medical imaging diagnosis and clinical decision support, aiming to standardize diagnostic reports, alleviate the shortage of clinical resources, and enhance telemedicine efficiency.

## Data Availability

All data produced in the present study are available upon reasonable request to the authors

## Key Points

### Question

Can a multimodal artificial intelligence (AI) system effectively generate diagnostic reports and provide interactive clinical decision support from ophthalmic B-scan ultrasound images?

### Findings

In this retrospective study of 54,696 ophthalmic B-scan ultrasound images and 9,392 reports from 31,943 patients, the OphthUS-GPT system achieved high performance in diagnostic report generation (ROUGE-L: 0.6131, CIDEr: 0.9818) and disease classification (>90% accuracy for common conditions). The integrated DeepSeek-R1-Distill-Llama-8B (DeepSeek) question-answering module showed excellent performance in accuracy, completeness, potential harm, and satisfaction, comparable to GPT4o and OpenAI-o1, and significantly outperforming other models.

### Meaning

These results demonstrate that the OphthUS-GPT multimodal system we developed is capable of automating report generation and enabling intelligent Q&A based on ophthalmic B-scan images, offering a novel solution for medical imaging diagnosis and clinical decision support.

## Author Contributions

Li, Yu, Zhang, Xu, Cheng.

Drafting of the manuscript: Gan, Chen, You.

Critical review of the manuscript for important intellectual content: All authors.

Statistical analysis: Chen, Qin, Han, Long.

Obtained funding: You.

Administrative, technical, or material support: Fan, Xin. Li, Yu, Zhang, Xu, Shao, Xiao. Li, Wan, T.

